# The characteristics and risk factors of burnout for public servants in North China

**DOI:** 10.1101/2024.05.24.24307862

**Authors:** Zhiyi Jia, Jinping Chen, Yalong Dang

## Abstract

**Objective:** Burnout significantly affected the working efficiency of public servants. This study aimed to identify the characteristics and risk factors of burnout among public servants in North China.

**Methods:** A cross-sectional online and anonymous survey was conducted between March 2024 and April 2024. The inclusion criteria included active public servants aged between 18 and 60 years with fluency in Chinese. Participants with mental disorders were excluded. The study utilized the Maslach Burnout Inventory-General Survey with Chinese adaptation to measure burnout across three dimensions: exhaustion, cynicism, and professional efficacy. The survey consisted of 16 seven-point Likert scale questions, with response options ranging from “never” to “daily,” and scored from 1 to 7. Participants with burnout had to meet at least one of the following criteria: 1) scoring in the upper third for exhaustion, 2) scoring in the upper third for cynicism, or 3) scoring in the lower third for professional efficacy. Severe burnout was defined as meeting all three criteria, moderate burnout was defined as meeting two criteria, and mild burnout was defined as meeting one criterion.

**Results:** The study included 1064 participants from seventeen provinces in North China. The breakdown of burnout levels among the participants was as follows: 34.3% (365/1064) experienced no burnout, 29.5% (314/1064) had mild burnout, 35.6% (379/1064) had moderate burnout, and 0.6% (6/1064) experienced severe burnout. The study revealed no significant differences in burnout risk based on gender, age, academic degrees, occupation types, and length of service. However, significant variations were observed in burnout risk based on job position, salary, income satisfaction, job security, work stress, and interpersonal ability.

**Conclusion:** In North China, two-thirds of public servants experienced occupational burnout. Job position, salary, income satisfaction, job security, work stress, and interpersonal ability were identified as the risk factors.

## Introduction

China is the second-largest economy globally, trailing the United States. It is classified as an upper-middle-income country by the World Bank, basing on China’s Gross National Income per capita[1]. While China has made significant strides in poverty reduction and economic growth since its reform and opening up in 1978, it still faces challenges in terms of income inequality[2] and regional disparities[3]. China’s economic growth is slowing down in the 2020s as it deals with a range of challenges from a rapidly aging population[4], higher unemployment[5].

The public sector, including state-owned enterprises, schools and hospitals, plays a central role in China’s economy[6]. Burnout is a state of emotional, physical, and mental exhaustion caused by prolonged or excessive stress[7]. Burnout is an influencing factor for the working efficiency of public servants, thus highly related to the efficacy of the public sector[8]. Recently, the burnout of public servants in China has become a growing concern[9], and while there is increasing awareness of the issue, identifying specific risk factors can be complex due to the following reasons: 1) While some studies have examined burnout among Chinese public servants, research in this area is still relatively limited compared to other countries[10]. 2) Cultural factors: The Chinese workplace culture and public service environment are distinct from those in Western countries[11]. Factors like hierarchical structures, performance pressures, and societal expectations can contribute to burnout in ways that might not be fully captured by existing research frameworks. 3) Data Collection Challenges: gathering reliable data on burnout among public servants can be challenging due to potential stigma and reluctance to report mental health issues. 3) Evolution of the Public Sector: China’s public sector is undergoing significant changes with reforms, modernization efforts, and increasing demands on public services[12].

This study aims to effectively address the characteristics and risk factors of burnout among Chinese public servants. This will enable the development of tailored interventions and policies to prevent and mitigate burnout in the public sector, and thus increase the working efficiency.

## Methods

### 2.1 Study design

This is a cross-sectional online and anonymous survey which was approved by the Institutional Review Board of Sanmenxia Central Hospital in accordance with the Declaration of Helsinki (reference No.20240319). All participants signed the consent documents and were enrolled between March 2024 and April 2024. Since we assessed the burnout of the participants, a physical address of help service and cellphone number were also provided in case of any consultant needed.

### 2.2 Inclusion and exclusion criteria

The inclusion criteria were active public servants at the age between 18 to 60 years and Chinese fluency. The participants with mental disorder were excluded.

### 2.3 Survey design and development

Maslach Burnout Inventory-General Survey (MBI-GS) originally developed by Christina Maslach et al[13] was used in this study with Chinese adaptation by Chaoping Li et al[14].

The process for adapting the Maslach Burnout Inventory-General Survey (MBI-GS) for use in China involved several steps[14]. Firstly, the questionnaire was independently translated into Chinese by four experts. Following this, six employees from diverse educational and professional backgrounds filled in the questionnaire and provided feedback, leading to modifications in the text expressions to form the initial Chinese version. Subsequently, two English experts back-translated the Chinese questionnaire into English, and this version was reviewed and adjusted by Michael Leiter, one of the main developers of MBI-GS.

The survey included 12 demographic questions, covering gender, age, academic degrees, occupation types, length of service, job position, salary, satisfaction, mental health, job security, work stress, and interpersonal ability.

Burnout was measured across three dimensions: exhaustion, cynicism, and professional efficacy using 16 seven-point Likert scale questions, with response options ranging from “never” to “daily” and scored from 1 to 7. The participant with burnout should meet at least one of the following criteria: 1) scoring in the upper third for exhaustion, 2) scoring in the upper third for cynicism, or 3) scoring in the lower third for professional efficacy. Severe burnout was defined as meeting all three criteria, moderate burnout was defined as meeting two criteria, and mild burnout was defined as meeting one criterion.

### 2.4 Data collection

The survey was conducted, and data was collected via an online survey system (https://www.wjx.cn). The questionnaire could be accessed through https://www.wjx.cn/vm/QI9xLUI.aspx. The raw data file is available at https://docs.google.com/spreadsheets/d/1yf8xsx5TT84uWWX7e9C8Vtp7AlqUheCsddUKY36Ni5o/edit?usp=sharing

### 2.5 Statistically analysis

SPSS 26.0 (IBM, SPSS Inc. USA) was used for data analysis. The quantitative data were presented as mean ± standard deviation. Group differences were assessed using Student’s t-tests and one-way analysis of variance (ANOVA). Logistic regression was used to identify risk factors for burnout, with a significance level of p < 0.05 indicating statistical significance.

## Results

### 3.1 Demographic characteristics of participant

A total of 1064 participants from seventeen provinces in the North China were finally enrolled in the study. The characteristics of these participants were described in Table 1.

**Table 1.** Demographic characteristics of participant.

| Characteristic |  | *n* | Percentage |
| --- | --- | --- | --- |
| Gender | Male | 304 | 28.6% |
|  | Female | 760 | 71.4% |
| Age<br>(years) | 18 -35 | 640 | 60.2% |
|  | 36 -55 | 401 | 37.7% |
|  | 55 and above | 23 | 2.2% |
| **Academic degree** | Associate degree | 122 | 11.5% |
|  | Bachelor's degree | 834 | 78.4% |
|  | Master's degrees or above | 108 | 10.2% |
| **Occupation** | Universities | 2 | 0.2% |
|  | State-owned enterprises | 431 | 40.5% |
|  | Public institutions | 616 | 57.9% |
|  | Private enterprise | 14 | 1.3% |
|  | Government officer | 1 | 0.1% |
| **Length of service (years)** | >30 years | 60 | 5.6% |
|  | 20-30 years | 76 | 7.1% |
|  | 6-20 years | 651 | 61.2% |
|  | < 6 years | 277 | 26.0% |
| **Job position** | Intern | 51 | 4.8% |
|  | Regular employee | 865 | 81.3% |
|  | Middle-ranking employee | 145 | 13.6% |
|  | Senior-ranking employee | 3 | 0.3% |
| **Salary (CNY)** | 1800 or less | 38 | 3.6% |
|  | 1800-3000 | 317 | 29.8% |
|  | 3,000 -5,000 | 414 | 38.9% |
|  | 5,000 -10,000 | 269 | 25.3% |
|  | 10,000 -20,000 | 24 | 2.3% |
|  | 20,000 or more | 2 | 0.2% |
| **Income satisfaction** | Not satisfied | 246 | 23.1% |
|  | Very satisfied | 68 | 6.4% |
|  | Satisfy | 225 | 21.1% |
|  | Fair | 525 | 49.3% |
| **Job security** | Full of uncertain | 359 | 33.7% |
|  | Uncertain | 256 | 24.1% |
|  | Safe | 449 | 42.2% |
| **Work stress** | No stress | 138 | 13.0% |
|  | Mild stress | 415 | 39.0% |
|  | Moderate stress | 318 | 29.9% |
|  | A lot of stress | 193 | 18.1% |
| **Interpersonal ability** | Good | 761 | 71.5% |
|  | mild social impairment | 251 | 23.6% |
|  | moderate social impairment | 41 | 3.9% |
|  | severe social impairment | 11 | 1.0% |

71.4% of participants were female, while males accounted for 28.6% in this survey. In terms of age, 640 participants (60.2%) were aged 18 to 35, 401 participants (37.7%) were aged 36 to 55, and 23 participants (2.2%) were over 55. The majority of participants held a bachelor’s degree (78.4%), followed by an associate degree (11.5%), and a master’s degree or higher (10.2%). The majority of participants worked in public institutions (57.9%), followed by state-owned enterprises (40.5%), with smaller numbers working in universities (0.2%), private enterprises (1.3%), or as government officers (0.1%). 651 participants (61.2%) had 6-20 years of service, 277 participants (26.0%) had less than 6 years of experience, 76 participants (7.1%) had 20-30 years of experience, and 60 participants (5.6%) had over 30 years of experience. 81.3% of participants were regular employees, 13.6% were middle-ranking employees, 4.8% were interns, and 0.3% were senior-ranking employees. Income distribution was as follows: 38 participants (3.6%) earned below 1,800 CNY, 317 participants (29.8%) earned 1,800 CNY to 3,000 CNY, 414 participants (38.9%) earned 3,000 to 5,000 CNY, 269 participants (25.3%) earned 5,000 CNY to 10,000 CNY, and 24 participants (2.3%) earned 10,000 CNY to 20,000 CNY. Only 2 participants (0.2%) earned over 20,000 CNY. Regarding “income satisfaction”, 49.3% of participants felt “fair”, 23.1% were not satisfied, 6.4% were “very satisfied”, and 21.1% were in the “satisfy” group. 42.2% of participants felt their job was “safe”, 33.7% felt it was “full of uncertain”, and 24.1% felt it was “uncertain”. Work stress levels varied with 39.0% experiencing mild stress, 29.9% experiencing moderate stress, 18.1% experiencing a lot of stress, and 13.0% experiencing no stress. 71.5% of participants felt good about their interpersonal abilities, while 23.6% reported mild social impairment, 3.9% reported moderate social impairment, and 1.0% reported severe social impairment.

### 3.2 Assessment of burnout in the three dimensions

The average score for burnout across three dimensions (exhaustion, cynicism, and professional efficacy) were 15.29±8.48, 11.85±6.62, and 29.89±9.17, respectively. Based on the burnout definition outlined in the Methods section, the cutoff values for exhaustion and cynicism were 12 or below, while for professional efficacy it was 14 or above (Table 2).

**Table 2.** The distribution of the three dimensions of burnout.

| Dimensions | Score | Cutoff value | Positive | Negative |
| --- | --- | --- | --- | --- |
| Exhaustion | $15.29 \pm 8.48$ | 12 | 581 | 483 |
| Cynicism | $11.85 \pm 6.62$ | 12 | 424 | 640 |
| Professional efficacy | $29.89 \pm 9.17$ | 14 | 85 | 979 |

Among the total of 1064 participants, 34.3% (365/1064) had no burnout, 29.5% (314/1064) had mild burnout, 35.6% (379/1064) had moderate burnout, and 0.6% (6/1064) suffered from severe burnout (Table 3).

**Table 3.** Severity of burnout.

| Severity of burnout | Numbers of participant | % |
| --- | --- | --- |
| No burnout | 365 | 34.3 |
| Mild burnout | 314 | 29.5 |
| Moderate burnout | 379 | 35.6 |
| Severe burnout | 6 | 0.6 |

### 3.3 Logistic regression of risk factors for influencing burnout

Taking burnout as the dependent variable (the absence of burnout is assigned as “0”, and the presence of burnout is assigned as “1”), gender, age, academic degrees, occupation types, length of service, job position, salary, income satisfaction, mental health, job security, work stress, and interpersonal ability were included in the logistic regression model. The total fitting information likelihood ratio test showed that the significance was p<0.01, indicating that the regression analysis was overall meaningful.

The results indicated no significant difference in the risk of burnout among the five factors including gender, age, academic degrees, occupation types, and length of service. However, significant differences were observed in the risk of burnout among six factors: job position, salary, income satisfaction, job security, work stress, and interpersonal ability. Further analysis of the six factors with significant differences is presented in Table 4:

**Table 4.** Logistic regression analysis of risk factors influencing burnout.

| Factor |  | OR | 95% CI | P value |
| --- | --- | --- | --- | --- |
| Gender | Male | 1.00 | - | - |
|  | Female | 1.09 | 0.751-1.576 | 0.655 |
| Age (years) | 18 -35 | 1.00 | - | - |
|  | 36 -55 | 0.78 | 0.512-1.173 | 0.228 |
|  | 55 and above | 1.13 | 0.296-4.330 | 0.857 |
| Academic degree | Associate degree | 0.89 | 0.538-1.470 | 0.646 |
|  | Bachelor's degree | 1.00 | - | - |
|  | Master or above | 0.89 | 0.514-1.525 | 0.662 |
| Occupation | Universities | 1.00 | - | - |
|  | State-owned enterprises | 0.00 | 0.00-0.00 | 0.999 |
|  | Public institutions | 0.00 | 0.00-0.00 | 0.999 |
|  | Private enterprise | 0.00 | 0.00-0.00 | 1.000 |
|  | Government officer | 0.24 | 0.00-0.00 | 1.000 |
| **Length of service** | >30 years | 0.56 | 0.217-1.43 | 0.224 |
|  | 20-30 years | 1.00 | - | - |
|  | 6-20 years | 0.84 | 0.438-1.597 | 0.589 |
|  | < 6 years | 0.93 | 0.426-2.043 | 0.862 |
| **Job position** | Middle-ranking employee | 1.00 | - | - |
|  | Senior-ranking employee | 0.12 | 0.007-1.965 | 0.138 |
|  | Regular employee | 2.32 | 1.317-4.08 | 0.004 |
|  | Intern employee | 2.75 | 1.058-7.172 | 0.038 |
| **Salary (CNY)** | 10,000 -20,000 | 1.00 | - | - |
|  | 1800-3000 | 0.19 | 0.053-0.68 | 0.011 |
|  | 1800 or less | 0.18 | 0.037-0.861 | 0.032 |
|  | 20,000 or more | 0.00 | 0.00-0.00 | 0.999 |
|  | 3,000 -5,000 | 0.23 | 0.068-0.779 | 0.018 |
|  | 5,000 -10,000 | 0.26 | 0.082-0.828 | 0.023 |
| **Income satisfaction** | Not satisfied | 1.00 | - | - |
|  | Very satisfied | 0.22 | 0.094-0.508 | <0.001 |
|  | Satisfy | 0.40 | 0.221-0.727 | 0.003 |
|  | Fair | 0.66 | 0.401-1.084 | 0.101 |
| **Job security** | Full of uncertain | 1.00 | - | - |
|  | Safe | 0.37 | 0.241-0.573 | <0.001 |
|  | Uncertain | 0.68 | 0.426-1.088 | 0.108 |
| **Work stress** | Moderate stress | 1.00 | - | - |
|  | Mild stress | 0.33 | 0.224-0.5 | <0.001 |
|  | No stress | 0.25 | 0.141-0.44 | <0.001 |
|  | A lot of stress | 2.09 | 1.096-3.991 | 0.025 |
| **Interpersonal ability** | Mild social impairment | 1.00 | - | - |
|  | Good | 0.34 | 0.213-0.53 | <0.001 |
|  | Moderate social impairment | 1.01 | 0.273-3.723 | 0.990 |
|  | Severe social impairment | 56563772.29 | 0.00-0.00 | 0.999 |

1. Job position:Relative to middle-ranking employees, the burnout of intern staff was 1.75 times higher, and the burnout of regular employees was 1.32 times higher. There was no significant difference in burnout risk between middle-ranking employees and senior-ranking employees.
2. Salary: The burnout of people with an income of less than 1,800 CNY was 82% higher, and the burnout of people with incomes between 1,800 to 3,000 CNY was 81% higher, and the burnout of people with incomes between 3,000 to 5,000 CNY was 77% higher compared to those with incomes of 10,000 to 20,000 CNY. There was no significant difference between people with an income of more than 20,000 CNY and those with an income of 10,000 to 20,000 CNY.
3. Income satisfaction: People who were “very satisfied” with their income had a 78% lower burnout than those who were dissatisfied with their income. People who were “satisfied” with their income had a 60% lower burnout than those who were dissatisfied with their income. People who felt “fair” with their income were generally the same as people who were dissatisfied with their income.
4. Job security: People who felt secure about their jobs had a 63% reduction in burnout compared to people who were uncertain about their job development. People who felt uncertain had a 32% reduction in job burnout compared to people who were “full of uncertainty” about their job development.
5. Work stress: People who experienced “a lot of stress” at work had 1.09 times higher burnout than those who felt “moderate stress”. People who felt “mild stress” at work had a 67% lower degree of burnout than those who felt “moderate stress”. People who felt “no stress” at work had a 75% lower degree of burnout than those who felt “moderate stress”. The greater the overall stress, the greater the burnout.
6. Interpersonal ability: People with no barriers to interpersonal communication had about 66% lower burnout than those with mild social impairment. There was no significant difference in burnout risk between people with moderate social impairment and mild social impairment. People with severe social impairment had no significant difference in burnout risk compared with those with mild social impairment.

## Discussion

While we did not have a definitive percentage of burnout across the entire Chinese population, studies suggested it was a significant issue, especially in certain professions[15,16]. In our study, the overall percentage of burnout for public servants was above 65%, which was lower than that of 72.9% in medical professionals[16], especially in the category of severe burnout (0.6% in our study versus 20.5% in Gao et al.’s study[16]). Our studies also suggested that job position, salary, income satisfaction, job security, work stress, and interpersonal ability were six independent risk factors for burnout. This might have been the first study describing the characteristics and risk factors of burnout for public servants in North China. These findings show that burnout was a serious concern in North China, particularly for those who held lower job positions, received lower salaries and income satisfaction, had lower job security and interpersonal ability, and experienced high work stress. It emphasized the need for further research and initiatives to promote well-being at work.

In our study, we found that intern staff and regular employees had 1.75 times and 1.32 times higher burnout than that of middle-ranking employees, indicating a higher job position might have a lower incidence of burnout. That might have been particularly because of 1) Higher positions often come with greater control over decision-making, work scheduling, and the ability to influence workplace conditions. This enhanced autonomy could reduce stress and burnout[17–19]. 2) Senior roles generally offer better salaries, benefits, and recognition, which could increase feelings of value and reduce stressors associated with financial worries[20]. 3) Leadership positions might provide a greater sense of meaning and contribution, buffering against burnout[21,22].

As far as we know, China is home to the largest middle-class cohort worldwide. The definition of the middle class in China was approximately a monthly income of 10,000 CNY to 20,000 CNY[23], according to the National Bureau of Statistics. In this study, we found that the participants with lower monthly income always had a higher incidence of burnout. This was consistent with the findings of the American Psychological Association[24]. That might have been particularly because people with lower incomes may have less access to resources that could help prevent or mitigate burnout, such as mental health care[25], stress management programs[26], or wellness benefits[27].

Job security was a risk factor associated with burnout, according to our study. We found that the participants who felt secure about their jobs had a 63% reduction in burnout compared to those who were uncertain about their job development. Low job security might have caused the reduction of motivation and engagement for a job, leading to feelings of cynicism[28–30]. However, some studies suggested that excessive job security without sufficient challenges might have led to boredom and dissatisfaction, contributing to burnout[31–33]. Csikszentmihalyi introduced the concept of “flow,” a state of optimal experience characterized by complete absorption in a challenging and engaging activity. He argued that a lack of challenge and growth opportunities could have led to boredom and apathy, hindering personal growth and well-being[31]. Additionally, job security was also closely associated with work stress. High work stress could have reduced the job security by increasing the fear of job loss[34] and reducing psychological well-being, thus increasing the incidence of burnout[35,36].

Moreover, in this study, we also found that participants with no barriers to interpersonal communication had about 66% lower burnout than those with social impairment. This was consistent with Bakker et al[37] and Wharton[38] studies. Individuals with strong interpersonal skills were better equipped to navigate workplace challenges, resolve conflicts, and build supportive relationships with colleagues. These positive interactions could have fostered a sense of belonging and camaraderie, reducing feelings of isolation and stress that contributed to burnout.

This study had two limitations. 1) most of the participants were from public institutions (hospitals and primary or middle schools) and state-owned enterprises, accounting for 98.4% in total. This might have been because of the location of launching the survey, which might have led to research bias. 2) only three senior-ranking employees (0.3% in total) took part in this survey. This might have been due to the confidentiality concerns and lack of time.

In conclusion, in North China, two-thirds of public servants experienced occupational burnout. Job position, salary, income satisfaction, job security, work stress, and interpersonal ability were the risk factors.

## Ethics Approval and Consent to Participate

The study protocol was approved by the ethics committee of Sanmenxia Central Hospital (reference No.20240319).

## Research Involving Humans

The study protocol strictly adhered to the clinical research principles outlined in the Declaration of Helsinki.

## Consent for Publication

All patients provided written informed consent prior to inclusion in the study.

## Availability of Data and Materials

The data supporting the findings of the article is available at: https://docs.google.com/spreadsheets/d/1yf8xsx5TT84uWWX7e9C8Vtp7AlqUheCsddUKY36Ni5o/edit?usp=sharing.

## Acknowledgements

None.

